# CharMark: A Markov Approach to Linguistic Biomarkers in Dementia

**DOI:** 10.1101/2025.07.14.25331543

**Authors:** Kevin Mekulu, Faisal Aqlan, Hui Yang

## Abstract

Dementia, one of the most prevalent neurodegenerative diseases, affects millions worldwide. Understanding linguistic markers of dementia is crucial for elucidating how cognitive decline manifests in speech patterns. Current non-invasive assessments like the Montreal Cognitive Assessment (MoCA) and Saint Louis University Mental Status (SLUMS) tests rely on manual interpretation and often lack detailed linguistic insight. This paper introduces a first-of-its-kind interpretable artificial intelligence (IAI) framework, *CharMark*, which leverages first-order Markov Chain models to characterize language production at the character level. By computing steadystate probabilities of character transitions in speech transcripts from individuals with dementia and healthy controls, we uncover distinctive character-usage patterns. The space character ““, representing pauses, and letters such as “`n`” and “`i`” showed statistically significant differences between groups. Principal Component Analysis (PCA) revealed natural clustering aligned with cognitive status, while Kolmogorov-Smirnov tests confirmed distributional shifts. A Lasso Logistic Regression model further demonstrated that these character-level features possess strong discriminative potential. Our primary contribution is the identification and characterization of candidate linguistic biomarkers of cognitive decline; features that are both interpretable and easily computable. These findings highlight the potential of character-level modeling as a lightweight, scalable strategy for early-stage dementia screening, particularly in settings where more complex or audio-dependent models may be impractical.

## 1 INTRODUCTION

Alzheimer’s Disease and related dementias (ADRD) represent a growing global health crisis. As of 2020, more than 55 million people worldwide were affected, and projections suggest this number will rise to 139 million by 2050 Alzheimer’s Disease International (2021). Early detection remains one of the most powerful levers for improving outcomes, yet current tools face fundamental challenges that limit their utility in real-world clinical practice.

Widely used screening tools, such as the Montreal Cognitive Assessment (MoCA) and the Saint Louis University Mental Status (SLUMS) exam, remain the default in many healthcare settings. However, they present several well-documented limitations:

- **Subjectivity and Interpretation Bias:** Outcomes can vary significantly depending on the examiner’s training, experience, and cultural background Karimi et al. (2022).
- **Time and Resource Burden:** These tools require clinician involvement and are time-consuming to administer and score De Roeck et al. (2019).
- **Limited Diagnostic Confidence:** Surveys reveal that as many as 40% of primary care providers lack confidence when diagnosing dementia based solely on these screenings Tsoi et al. (2015).
- **Educational and Cultural Bias:** Performance on these tests can be influenced by a patient’s education level and language proficiency, increasing the risk of misdiagnosis Tumas et al. (2016).

Given these constraints, there is an urgent need for more objective, efficient, and culturally adaptable screening methods that can be deployed at scale.

This study introduces a novel modeling framework—*CharMark*—designed to uncover early linguistic biomarkers of dementia through character-level analysis of speech transcripts. Figure 1 presents a graphical depiction of the Markov network at the heart of our approach, constructed from transitions between characters in recorded speech.

**Figure 1.**
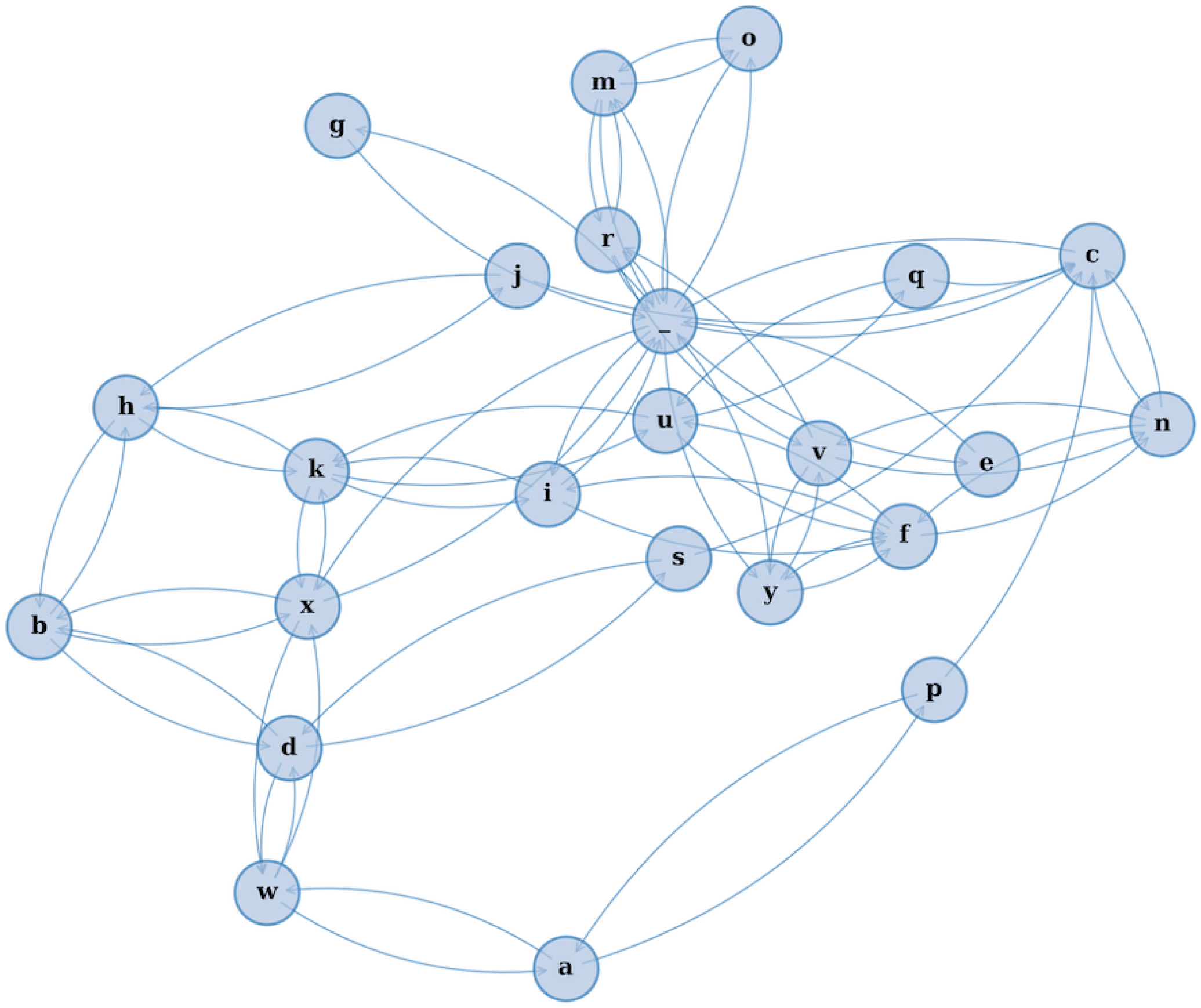
A visual rendering of the character-level Markov Chain model used in our study. Each node represents a character, and directed edges illustrate the probability of transitioning from one character to another based on observed speech transcripts. This network reveals the structural “fingerprint” of how language flows — capturing micro-level speech dynamics often imperceptible to human raters.

Unlike prior models that emphasize high-level semantic or acoustic features, we analyze language at its most granular level: transitions between individual characters. Our framework computes **steady-state probabilities** for each character using a first-order Markov Chain model Hayes (2013), which allows us to quantify how often each character appears in the long-run behavior of speech. This yields interpretable linguistic fingerprints that are not only compact but also agnostic to language and speaker variability.

To explore whether these fingerprints contain clinically meaningful information, we perform unsupervised **k-means clustering** and visualize group separation via **Principal Component Analysis (PCA)**. We also apply the **Kolmogorov-Smirnov (KS)** test Massey Jr (1951) to statistically confirm character-level differences between dementia and control groups. Finally, we use a **Lasso Logistic Regression** model to assess the discriminative value of these features, while maintaining interpretability.

Our contributions are as follows:

- **A novel character-level approach to linguistic biomarker discovery:** We introduce an interpretable AI framework based on steady-state character probabilities derived from first-order Markov Chains.
- **Identification of distinctive linguistic signals:** Our analysis highlights specific characters—such as the space character (indicating pauses), “`n`”, and “`i`” that show statistically significant differences in usage between groups.
- **Validation using transparent statistical tools:** Through clustering, hypothesis testing, and logistic regression, we confirm the potential of these features as early-stage indicators of cognitive decline.

This study begins by reviewing prior research on linguistic analysis in Alzheimer’s Disease and Related Dementias (ADRD), establishing the context for our work. We then describe our methodology, including the dataset, preprocessing steps, and the implementation of our character-level Markov model. Our focus on character-level transitions rather than words or sentences, stems from the hypothesis that microstructural disruptions in language (such as altered pause patterns or character repetition) may serve as early markers of cognitive impairment. By operating at this fine-grained level, our framework captures subtle changes in language production that often precede higher-level semantic breakdowns. The remainder of the paper presents our results, interprets the identified linguistic signals, and discusses their implications for biomarker discovery and future screening applications.

## 2 MATERIALS AND METHODS

### 2.1 Overview of the Approach

This study introduces an interpretable modeling framework—CharMark—for identifying early linguistic biomarkers of dementia from transcribed speech. Our approach treats each transcript as a character sequence and models it using a first-order Markov Chain. From this, we compute a 27-dimensional vector representing the steady-state probability of each character (a–z and the space character). These vectors are then analyzed through clustering, dimensionality reduction, hypothesis testing, and logistic regression to evaluate group-level differences and the potential diagnostic value of the extracted features.

Figure 2 provides a conceptual overview of the CharMark pipeline.

**Figure 2.**
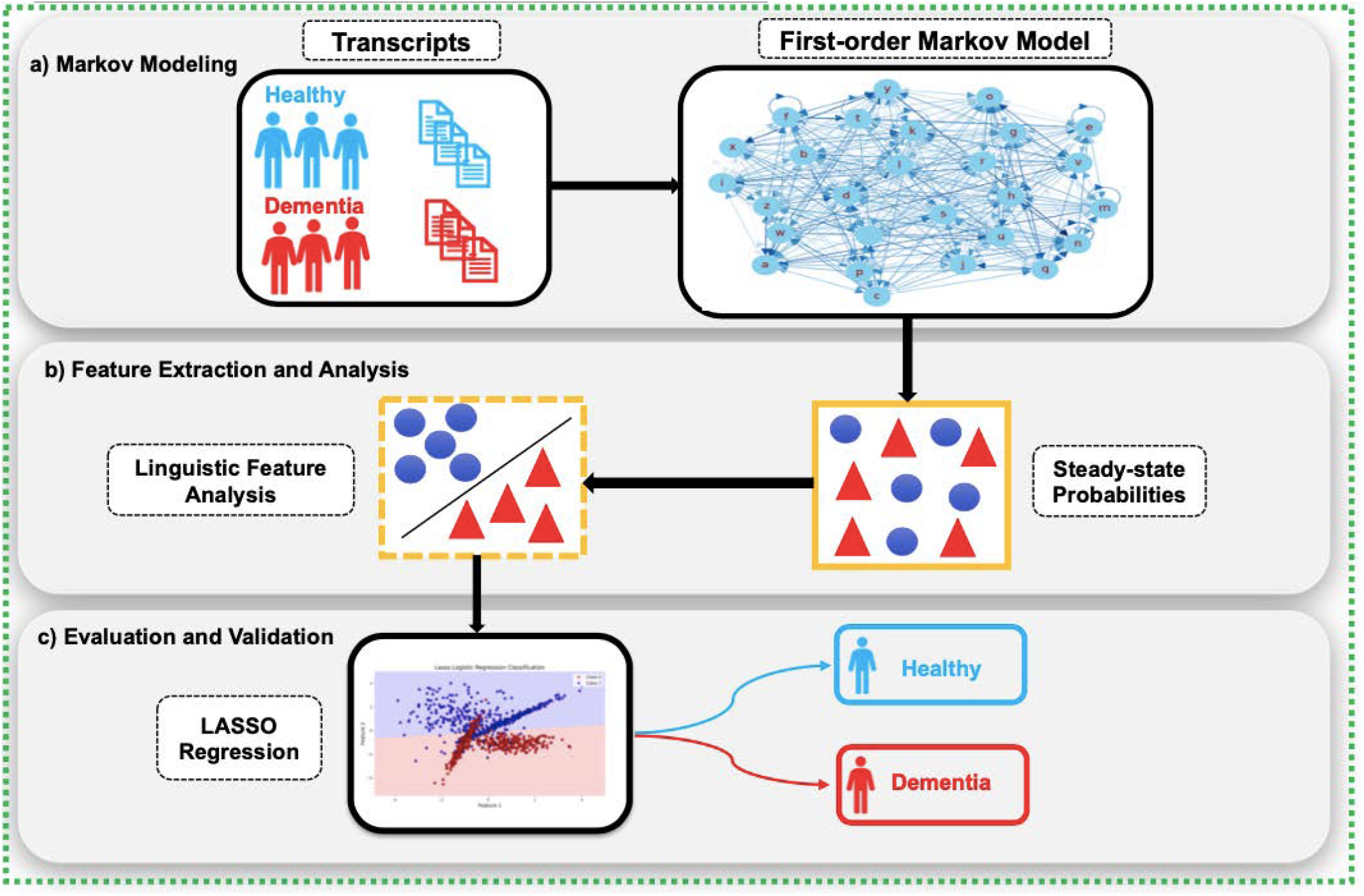
The CharMark framework begins with transcript preprocessing, followed by character-level Markov modeling to extract steady-state distributions. These linguistic fingerprints are then analyzed using statistical and machine learning techniques to identify and validate early cognitive biomarkers.

### 2.2 Dataset and Preprocessing

We used transcripts from the DementiaBank Pitt Corpus, a widely cited dataset in dementia and speech-language research. All participants were asked to describe the Boston Cookie Theft picture, a standardized elicitation task designed to generate naturalistic but structured language samples Goodglass et al. (2001).

The dataset included:

- 310 transcripts from 168 participants with Alzheimer’s Disease (AD)
- 242 transcripts from 98 cognitively healthy controls

Each transcript was lowercased, and non-alphabetic characters were removed. We retained the space character to preserve pausing patterns. The result was a discrete sequence of characters over a 27-character vocabulary (26 letters plus space), which served as the basis for Markov modeling.

### 2.3 Markov Chain Modeling

Markov Chain modeling offers a mathematically grounded and interpretable framework for capturing sequential dependencies in language. Its ability to model transition dynamics with minimal assumptions makes it well suited for identifying subtle patterns that may serve as early digital biomarkers Ross (2014). Building on our prior work using symbolic recurrence and character-level modeling to capture linguistic markers of cognitive decline Mekulu et al. (2025b,a), we adopt a first-order Markov Chain approach in this study.

Each transcript was modeled as a first-order Markov Chain, where the probability of observing character *s*_*j*_ depends only on the immediately preceding character *s*_*i*_. Transition probabilities were estimated using smoothed frequency counts with Laplace smoothing (*α* = 0.01), a small constant selected to mitigate zero-probability transitions while preserving the sparsity of natural language character distributions:

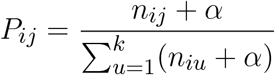

where *n*_*ij*_ is the number of observed transitions from character *s*_*i*_ to *s*_*j*_, and *k* = 27.

To derive the long-run behavior of character usage, we computed the steady-state distribution *π* by solving the eigenvector equation:

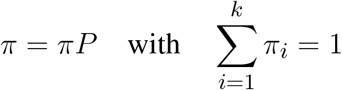

This steady-state vector, which reflects the long-term usage frequency of each character under the Markov process, was extracted for every transcript, forming a 552 × 27 feature matrix. These steady-state distributions serve as compact, interpretable fingerprints of linguistic structure across groups.

### 2.4 Clustering and Dimensionality Reduction

We applied *k*-means clustering to the steady-state vectors to explore group separability in an unsupervised setting. The optimal number of clusters was determined using silhouette analysis, which identified *k* = 2 as the most stable solution corresponding closely to the ground truth diagnostic labels of healthy controls and dementia patients. This finding suggests that meaningful cognitive signals are embedded in the character-level structure of language, independent of any supervised learning.

To better understand the geometry of these features, we performed Principal Component Analysis (PCA) on the 27-dimensional steady-state vectors. PCA reduces dimensionality while preserving the directions of greatest variance, enabling clearer visual interpretation. As shown in Figure 3, the resulting projection reveals a striking separation between clusters, with transcripts from dementia subjects forming a more compact and shifted group compared to the wider distribution of controls. This emergent clustering reinforces the hypothesis that cognitive decline alters linguistic dynamics in ways that are both measurable and visually discernible at the character level.

**Figure 3.**
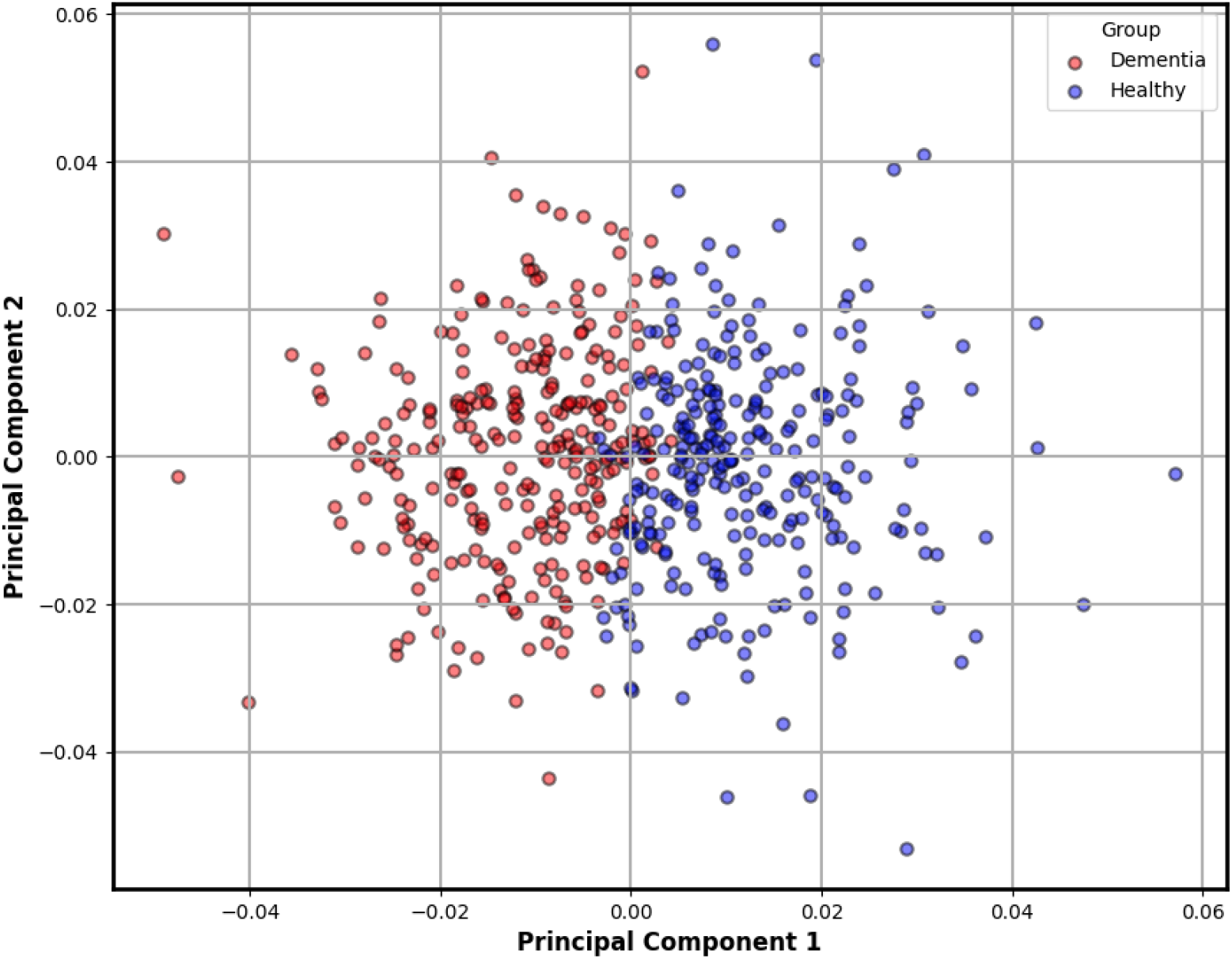
Two-dimensional PCA projection of steady-state character distributions derived from each transcript. This plot reveals a natural separation between dementia and control groups, as indicated by unsupervised *k*-means clustering. Transcripts from dementia subjects (red) tend to form a tighter, shifted cluster, suggesting reduced linguistic variability compared to the broader distribution of healthy controls (blue). This emergent structure reinforces the potential of character-level dynamics as interpretable biomarkers of cognitive decline.

### 2.5 Statistical Testing of Character Features

To identify which characters contributed most to group-level differences, we conducted two-sample Kolmogorov-Smirnov (KS) tests on each of the 27 character distributions (dementia vs. control). The KS statistic quantifies the maximal distance between the empirical cumulative distribution functions:

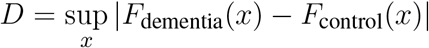

We selected the KS test due to its non-parametric nature and sensitivity to both location and shape differences between distributions, making it appropriate for features that may not follow Gaussian assumptions. Significance was evaluated at *α* = 0.05 with Bonferroni correction to control for multiple comparisons across the full character set. This conservative adjustment reduces false positives, ensuring that only robust distributional shifts are flagged as candidate biomarkers.

### 2.6 Validation via Lasso Logistic Regression

To assess the predictive relevance of the steady-state features, we trained a Lasso Logistic Regression model using the 27 character probabilities. Lasso was chosen for its ability to perform both classification and feature selection, enabling a sparse and interpretable solution. We evaluated model performance using the area under the Receiver Operating Characteristic curve (ROC-AUC).

### 2.7 Justification for Transcript-Based Character Modeling

While many dementia studies utilize acoustic features, we focused on text-based analysis to enhance interpretability, reproducibility, and cross-linguistic applicability. Character-level modeling captures finegrained language disruptions such as excessive pausing, rigid phrasing, and letter-specific anomalies that are often masked in higher-level or audio-driven analyses. Moreover, this approach is computationally efficient, robust to noise, and aligned with emerging needs for lightweight, privacy-preserving digital biomarkers in remote or resource-constrained environments.

## 3 RESULTS

### 3.1 Character-Level Distributions Reveal Salient Differences

We first examined steady-state character distributions to identify linguistic signals that differed significantly between groups. The Kolmogorov-Smirnov (KS) test revealed that several characters, including the space character ““, “`n`”, and “`i`”, showed statistically significant distributional shifts between dementia and control groups (Bonferroni-corrected *p <* 0.05). These characters reflect changes in pacing, repetition, and lexical structure often observed in cognitive decline. Table 1 summarizes the top-ranked features based on KS statistic.

**Table 1.** Kolmogorov-Smirnov test results for steady-state probabilities of selected characters. Significant differences were observed between dementia and control groups, with the space character showing the strongest effect.

| Character | KS Statistic | P-Value |
| --- | --- | --- |
| (space “ ”) | 0.286 | $2.50 \times 10^{-10}$ |
| n | 0.193 | $6.23 \times 10^{-5}$ |
| i | 0.192 | $7.13 \times 10^{-5}$ |
| c | 0.184 | $1.71 \times 10^{-4}$ |
| h | 0.181 | $2.19 \times 10^{-4}$ |

### 3.2 Space Character as a Primary Marker of Pausing Behavior

Among all features, the space character emerged as the most distinctive. Figure 4 visualizes the empirical distribution of steady-state space probabilities across groups. Dementia transcripts exhibit a rightward shift in the distribution, reflecting longer or more frequent pauses; likely tied to disrupted fluency.

**Figure 4.**
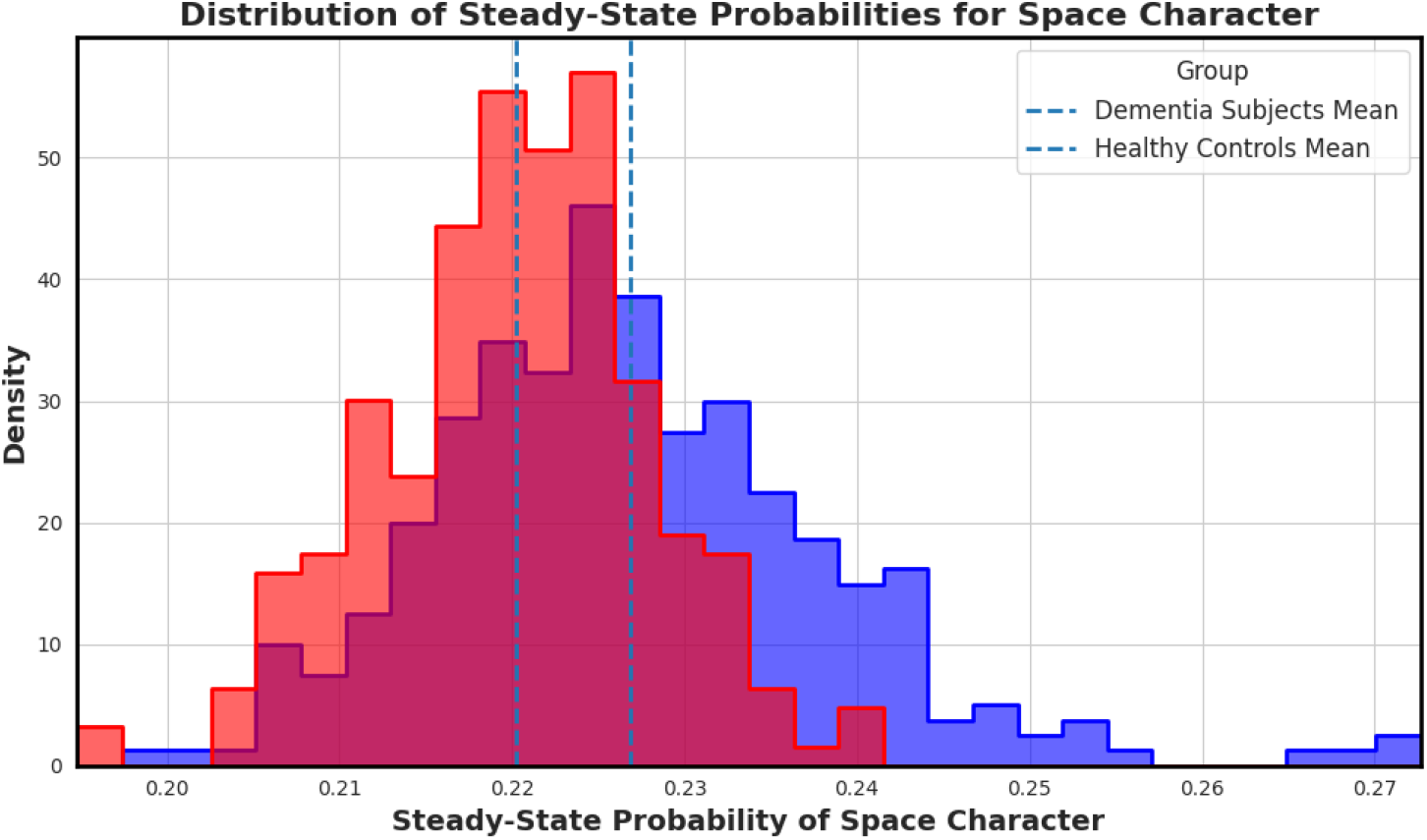
Distribution of steady-state probabilities for the space character across transcripts. Transcripts from individuals with dementia (red) show a clear shift in mean compared to controls (blue), highlighting increased or prolonged pauses that may reflect speech hesitancy or disrupted fluency.

To further probe this finding, we plotted a rolling mean of the space character probability across transcript indices (Figure 5). While average values remain elevated in dementia subjects, healthy controls display greater variance, potentially reflecting more adaptive or dynamic speech rhythm. The convergence of elevated mean and lower variance in dementia aligns with prior hypotheses about pacing rigidity in cognitive decline.

**Figure 5.**
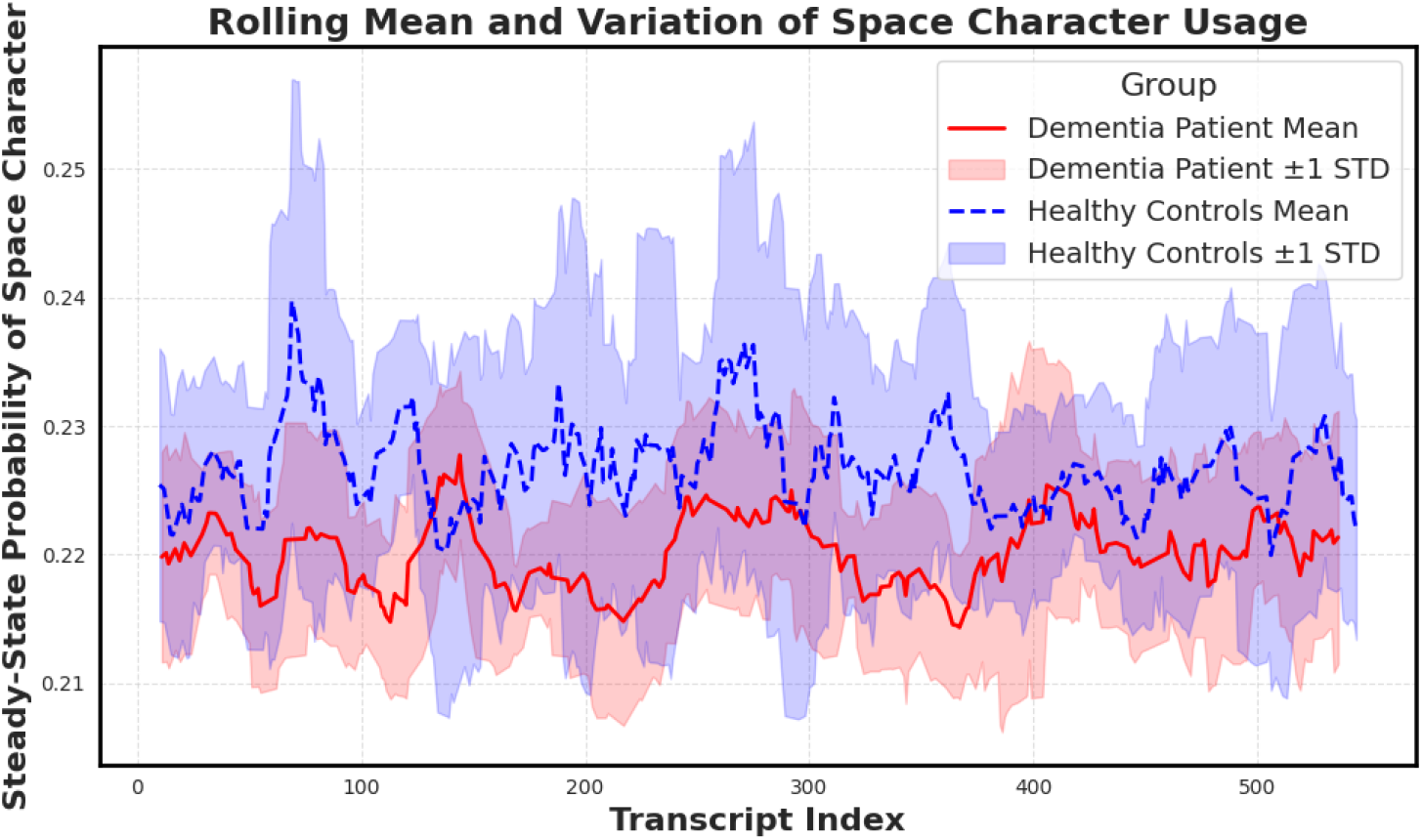
Rolling mean and standard deviation of the space character’s steady-state probability across transcripts. Dementia subjects show elevated means and notably reduced variance, supporting the hypothesis that cognitive decline manifests in more rigid and less adaptive pausing behavior.

### 3.3 Validation through Lasso Logistic Regression

To evaluate the diagnostic potential of our extracted features, we trained a Lasso Logistic Regression model using the 27-dimensional steady-state character vectors. The model was trained in a binary classification setting (dementia vs. control), and the resulting Receiver Operating Characteristic (ROC) curve is shown in Figure 6. The model achieved an area under the curve (AUC) of 0.806, indicating strong discriminative performance from the character-level features alone.

**Figure 6.**
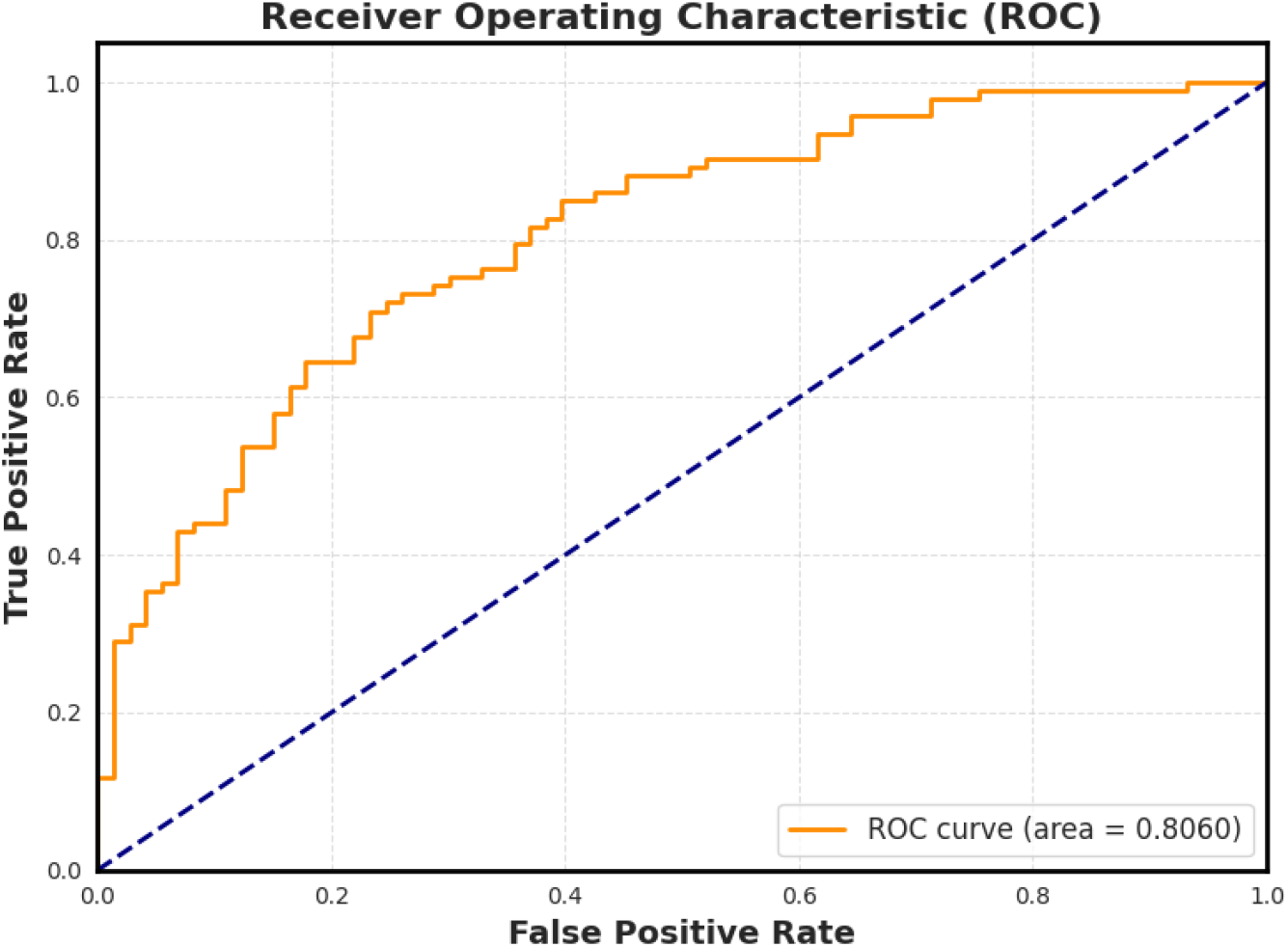
Receiver Operating Characteristic (ROC) curve for the Lasso Logistic Regression model trained on steady-state character features. The model achieves an AUC of 0.806, confirming that even low-level linguistic structures such as pause frequency and character usage carry sufficient signal to distinguish cognitive status.

Lasso regularization yielded a sparse solution, emphasizing only the most informative features. Notably, the space character remained among the most predictive features selected by the model, reinforcing its potential role as a candidate digital biomarker.

### 3.4 Structural Differences in Markov Networks

While scalar features like steady-state probabilities offer useful diagnostic signals, structural properties of the underlying transition networks also carry valuable insights into how linguistic rigidity manifests. To qualitatively explore this, we visualized first-order Markov transition networks from two representative transcripts, one from a dementia subject and one from a healthy control (Figure 7).

**Figure 7.**
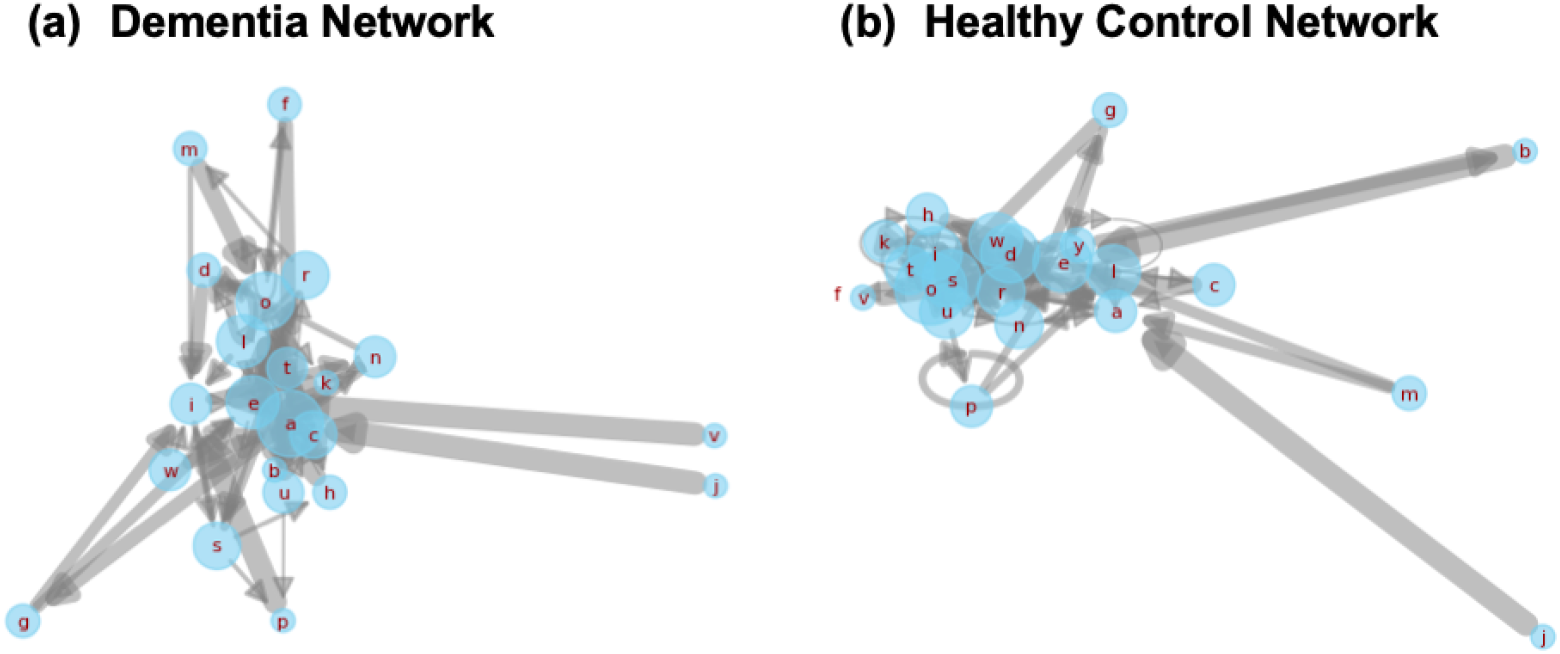
Side-by-side Markov transition networks generated from transcripts of a dementia subject and a healthy control. Each node corresponds to a character, and edge thickness represents transition probability. The dementia network (left) displays dense self-loops and concentrated local transitions, reflecting reduced lexical variety. The healthy network (right) reveals broader, more exploratory transitions, characteristic of flexible and adaptive language use.

The dementia network is dominated by self-loops and short-range transitions among a limited subset of characters, suggesting repetitive or constrained lexical production. In contrast, the control network exhibits greater transition diversity and broader edge connectivity, consistent with richer, more adaptive language use. These structural differences offer a complementary lens into how cognitive impairment alters the foundational dynamics of speech.

## 4 DISCUSSION

This study introduces a novel, interpretable framework, CharMark, for detecting linguistic signals of cognitive decline through character-level modeling of transcribed speech. By leveraging first-order Markov Chains and extracting steady-state probabilities, we uncover low-level features that capture subtle disruptions in language structure. These features, including the space character and letters such as “`n`” and “`i`”, exhibited significant distributional differences between individuals with dementia and healthy controls.

Our use of character-level modeling departs from the dominant paradigm in dementia speech analysis, which typically focuses on higher-level semantic or acoustic features. While these approaches have demonstrated predictive power, they often require large labeled datasets, complex tuning, and can be challenging to interpret in clinical settings. In contrast, CharMark produces compact, transparent features that offer a granular view of how cognitive impairment affects speech mechanics, making it particularly well suited for scalable and explainable biomarker discovery.

The elevated steady-state probability of the space character in dementia transcripts, combined with reduced variance, suggests a loss of dynamism in pause behavior. This aligns with clinical observations of reduced fluency, increased hesitation, and monotonic delivery in patients with early cognitive decline. Likewise, the reduced lexical diversity visualized through the Markov networks further supports the hypothesis that cognitive impairment constrains linguistic flexibility. Our findings thus reinforce the emerging view that early-stage dementia manifests not only in what is said, but how language is structured and delivered at a micro level.

Importantly, the Lasso Logistic Regression model yielded strong discriminative power (AUC = 0.806) using only 27 character-level features. The model’s sparsity highlights which transitions carry the most diagnostic value, offering a level of interpretability often missing from more complex black-box systems. This simplicity enhances trustworthiness and facilitates integration into clinical workflows, where transparent, actionable insights are essential.

These findings build upon and complement our prior work on symbolic recurrence analysis and character-level embeddings for cognitive assessment Mekulu et al. (2025b,a), while introducing a new perspective grounded in steady-state transition behavior. Compared to symbolic recurrence, the Markov modeling approach is computationally lightweight, more intuitive to interpret, and easier to deploy across language settings, particularly when acoustic data is unavailable or incomplete.

That said, several limitations merit discussion. First, the current analysis is based on a single structured elicitation task (the Cookie Theft description), which may limit generalizability to other speech contexts. Second, demographic variables such as age, education, and language background were not explicitly modeled, which could introduce subtle biases. Third, while steady-state probabilities offer a snapshot of character usage, they do not capture higher-order or context-dependent transitions. Future work could explore second-order models, recurrence dynamics, or integrate temporal attention mechanisms to enrich the representation.

Nonetheless, this work advances the field of digital biomarkers by highlighting the diagnostic potential of low-level, text-based features. By identifying character-level shifts that correspond to cognitive decline, CharMark contributes a novel layer of explainable signal that can enhance multimodal models and inform early-stage screening tools, especially in resource-constrained or multilingual settings where speech transcripts may be easier to collect than high-fidelity audio.

## 5 CONCLUSIONS

This study presents CharMark, a novel framework for identifying early-stage linguistic biomarkers of dementia using character-level Markov modeling. By analyzing steady-state transition probabilities in transcribed speech, we uncover compact, interpretable features that distinguish individuals with cognitive decline from healthy controls. Our results demonstrate that even the most granular units of language such as individual characters and pauses, encode meaningful diagnostic signal. CharMark offers a lightweight and transparent alternative to traditional semantic or acoustic approaches, making it especially well suited for scalable deployment in digital health tools. As speech-based screening continues to evolve, character-level modeling can serve as a critical foundation for interpretable and accessible cognitive assessment. Future work will extend this approach across diverse tasks and populations, and explore its integration into broader multimodal frameworks for precision brain health monitoring.

## Data Availability

The data that support the findings of this study are available from the Pitt Corpus within DementiaBank, maintained by TalkBank at Carnegie Mellon University and the University of Pittsburgh School of Medicine. Access to the data in DementiaBank is password protected and restricted to members of the DementiaBank consortium group. In accordance with TalkBank rules, any use of data from this corpus must be accompanied by appropriate corpus references and acknowledgment of grant support (NIA AG03705 and AG05133). Interested researchers can request access to the Pitt Corpus by visiting https://dementia.talkbank.org/access/English/Pitt.html. Established researchers and clinicians working with dementia can contact TalkBank to request access credentials.

## AUTHOR CONTRIBUTIONS

KM conceived the study, designed the methodology, performed the analysis, and drafted the manuscript. FA and HY supervised the project, contributed to the formulation of the methodological framework, and provided critical revisions. All authors reviewed and approved the final manuscript.

## ACKNOWLEDGMENTS

The authors of this work would like to acknowledge the NSF grants IIS-2302834 and MCB-1856132 for funding this research. Any opinions, findings, or conclusions found in this paper originate from the authors and do not necessarily reflect the views of the sponsor. The Pitt Corpus data used in this study was collected with support from NIA grants AG03705 and AG05133.

## SUPPLEMENTAL DATA

No supplemental data were generated or analyzed for this study.

